# The COVID HOME study research protocol: Prospective cohort study of non-hospitalised COVID-19 patients

**DOI:** 10.1101/2022.08.14.22278762

**Authors:** A. Tami, B.T.F. van der Gun, K.I. Wold, M.F. Vincenti-González, A.C.M. Veloo, M. Knoester, V.P.R. Harmsma, G.C. de Boer, A.L.W. Huckriede, D. Pantano, L. Gard, I. Rodenhuis-Zybert, V. Upasani, J. Smit, A. Dijkstra, J. de Haan, J. van Elst, J. van den Boogaard, S. O’ Boyle, L. Nacul, H.G.M. Niesters, A.W. Friedrich

## Abstract

**Background:** Guidelines on COVID-19 management are developed as we learn from this pandemic. However, most research has been done on hospitalised patients and the impact of the disease on non-hospitalised and their role in transmission are not yet well understood.

The COVID HOME study conducts research among COVID-19 patients and their family members who were not hospitalised during acute disease, to guide patient care and inform public health guidelines for infection prevention and control in the community and household.

**Methods:** An ongoing prospective longitudinal observational study of COVID-19 outpatients was established in March 2020 in the Netherlands. Laboratory confirmed SARS-CoV-2 infected individuals of all ages that did not merit hospitalisation, and their household (HH) members, were enrolled after written informed consent. Enrolled participants were visited at home within 48 hours after initial diagnosis, and then weekly on days 7, 14 and 21 to obtain clinical data, a blood sample for biochemical parameters/cytokines and serological determination; and a nasopharyngeal/throat swab plus urine, stool and sperm or vaginal secretion (if consenting) to test for SARS-CoV-2 by RT-PCR (viral shedding) and for viral culturing. Weekly nasopharyngeal/throat swabs and stool samples, plus a blood sample on days 0 and 21 were also taken from HH members to determine whether and when they became infected. All participants were invited to continue follow-up at 3-, 6-, 12- and 18-months post-infection to assess long-term sequelae and immunological status.

**Preliminary Results:** A total of 256 participants belonging to 103 HH were included of which, 190 (74.2%) were positive for SARS-CoV-2 infection. Most individuals (183/190, 96.3%) developed mild to moderate disease. At the time of writing, all participants had reached the 3 and 6 month time-points of the long-term follow-up, while approximately 78% reached 12 month and 23% the 18 month time-point. Preliminary analysis showed that 43% (52/121) positive individuals reported having complaints at 3 months post-infection, while 42.7% (61/143) had complaints at 6 months.

## Introduction

Knowledge on COVID-19, the disease caused by coronavirus SARS-CoV-2, is evolving at an enormous pace since the virus emerged in Wuhan, China at the end of 2019 [3, 4]. SARS-CoV-2 was first detected in the Netherlands at the end of February 2020 [5] and rapidly spread throughout the country causing a high mortality and morbidity in those infected during what will be later known as the first COVID-19 wave [6]. At the time and still nowadays, most of the research had been focussed on hospitalised infected individuals [7, 8], while much less is known about the acute and long-term clinical evolution in non-hospitalised patients. Moreover, the need to understand when individuals are not infectious any longer and can resume their working activities without the risk of transmitting the virus to others was an urgent question then and continues to be relevant given the emergence of new SARS-CoV-2 variants [9].

To answer these questions, the COVID HOME study (https://www.umcg.nl/-/covid-home) was set up in March 2020 by researchers from the Department of Medical Microbiology and Infection Prevention of the University Medical Center Groningen (UMCG), the Netherlands in collaboration with the Municipal Health Services (GGD in Dutch) of the north of the Netherlands. The project focusses on the impact of COVID-19 on non-hospitalised individuals, their acute and long-term clinical presentation, infectious period and transmission routes. We aimed to answer research questions that would inform public health guidelines on biosafety and transmission-control measures, as well as understand COVID-19 disease and its management in home-isolated patients.

Guidelines for quarantine and isolation measures vary by country and between regions within countries. Understanding the relevance of long-term (infectious) viral shedding in infected individuals [10] is key to determine when to de-escalate infection prevention and control practices and allow individuals to resume activities. Viral shedding in stool has proved relevant for monitoring the epidemic evolution through wastewater surveillance [11]. Moreover, the dynamics of respiratory and faecal viral shedding in the same individual may be linked to differing clinical presentations and may also give clues about a potential faecal-oral transmission route [12, 13]. Other pathways, such as via urine or sexual intercourse, may play a role in transmission [14]. However, it is still unclear how frequent and relevant is the presence of SARS-CoV-2 virus in vaginal and seminal fluids [15-17]. Furthermore, understanding SARS-CoV-2 transmission within restricted indoor areas, such as households (HH), gives detailed insight into transmission dynamics and transmission routes amongst contacts [18]. This may shed light on how HH transmission contributes to the overall community transmission (critical to sustain a pandemic), becoming a potential target for intervention.

Characterisation of the clinical phenotypes of patients with ‘mild’ disease is still lacking. The WHO clinical progression scale [19] classifies mild disease into asymptomatic and into two broad symptomatic strata. However, COVID-19 patients have diverse clinical presentations encompassing the number and duration of symptoms [20] as well as different alterations of laboratory parameters. Some of these clinical phenotypes could be linked to evolution to severe disease and/or to the development of long-term sequelae [21]. Several blood parameters including cytokines, have been identified as markers of COVID-19 disease severity [22, 23] while other studies have characterised blood parameters to predict COVID-19 diagnosis in patients presenting to hospital with symptoms [22, 24]. However, data collected in a systematic and longitudinal manner soon after infection will better characterise the clinical features and laboratory parameters (haematological, biochemical, immunological) in outpatient individuals convalescing at home and their potential as early markers of disease progression. A better understanding of the disease course in outpatients is needed to guide standard of care to properly manage patients at home, administer early treatment and predict progression to severe or long-term disease on time.

Here we report the design of a prospective observational cohort study of home-isolated SARS-CoV-2 infected individuals and their HH members. The main strength of this study is the early, systematic, prospective follow-up with an intense sampling/data collection of non-hospitalised COVID-19 patients during the acute disease evolution with a medium and long-term follow-up. The comprehensiveness, diverse data collection and analysis of the study described here will allow us and other researchers to add knowledge to the already ongoing fast research on COVID-19.

## Objectives

The objectives of the study have been divided into two work packages (WP) and their respective sub-objectives.

1. In WP1 we aim to measure the duration and routes of viral shedding, genetic diversity, and development of immunity of non-hospitalised individuals with COVID-19 to improve guidelines for biosafety and patient isolation by:
  1.1. Determining the viral load kinetics and duration of viral shedding of SARS-CoV-2 RNA in bodily specimens through real-time reverse transcription polymerase chain reaction (real-time RT-PCR)
  1.2. Evaluating the viability of the virus in different specimens and at various time points of patient follow-up, by viral culturing
  1.3. Determining the relation of viral load kinetics and the appearance and kinetics of anti-SARS-CoV-2 IgM, and IgG antibodies over time
  1.4. Determining the genetic diversity of SARS-CoV-2 through whole-genome sequencing (WGS)
  1.5. Determining the chain of transmission within the HH and into the working environment, especially for healthcare settings, through viral WGS
2. Within WP2 we expect to aid in establish guidelines for the management of (non-hospitalised) COVID-19 patients at home, including early detection of predicting factors for severity and long-term consequences by:
  2.1. Describing the clinical spectrum of COVID-19 in non-hospitalised individuals and determine potential risk factors for mild versus more severe clinical evolution
  2.2. Assessing the development of long-term sequelae and post-viral fatigue syndrome (PVFS) in non-hospitalised COVID-19 individuals
  2.3. Correlating laboratory parameters and cytokine changes with clinical evolution
  2.4. Determining the relation of viral loads & anti-SARS-CoV-2 antibody kinetics with clinical evolution

## Rationale

The data gathered in **WP1** will inform public health guidelines on biosafety, duration of home isolation and de-escalation measures as well as improve our understanding of transmission chains in the community (HH, workplace, healthcare centres). In **1.1** we aim to understand whether viral detection in different body specimens in non-hospitalised individuals differs from what is being found in hospitalised patients, thus understanding the contribution of outpatients to the overall transmission of COVID-19. Here, we tested for SARS-CoV-2 RNA on systematically and simultaneously collected clinical specimens from different body sites (respiratory, faeces, blood, urine, semen and vaginal fluid) for at least 4 weeks, giving detailed data to compare to the clinical evolution and other parameters such as immune response (**1.3**) and viral genotypes (**1.4**). In previous studies SARS-CoV-2 RNA was found in all of these specimens with conflicting results for vaginal fluid [15, 17]; with blood, urine and semen found positive in a much lower frequency than respiratory samples and faeces [14, 25, 26]. Secondly, in **1.2**, the RNA-positive specimens were cultured to ascertain if infectious virus could be found in the samples mentioned in **1.1**, at different time points during the clinical evolution. The presence and duration of infectious virus in different specimens influences infection prevention measures. Recent studies showed shedding of infectious virus in respiratory samples from critical hospitalised patients to last up to 20 days (median=8 days) since symptom onset [27], while other studies detected infectious virus in asymptomatic or mildly affected patients up to 10 days [10] but not after two weeks of diagnosis [28].

Follow-up studies on COVID-19 patients have shown that anti-SARS-CoV-2 IgM appears at around 4-14 days from symptom onset, while IgG seroconversion is seen between days 7-28 [29, 30]. These cohorts were mostly composed by hospitalised patients and IgG seroconversion was delayed in less severe cases [30], while mild SARS-CoV-2 infections induced a more modest antibody response [31]. **In 1.3** anti-SARS-CoV-2 antibody dynamics of our cohort of patients were measured and are being aligned with the viral shedding and virus culture results, to evaluate antibody kinetics, seroconversion rates and duration and the relation of serological response with viral clearance and virus viability. To assess long-term immunity, IgG data will be obtained up to 18 months post-infection in consenting patients (see Methods).

**In 1.4** we perform WGS analysis to determine the SARS-CoV-2 genetic diversity in the population under study. We want to establish the molecular and macro-epidemiologic context by comparing the sequences from participants with known sequences in our cross-border region as well as from international sequences to understand introduction and first seeding events of transmission into our region. WGS studies have shown that independent introductions resulted in patterns of local or national spread of viral types leading to lineage replacement [32, 33]. WGS was key to determine patterns of transmission among health care workers (HCW) in NL [34]. Transmission of SARS-CoV-2 within healthcare institutions seems to be one of the most important drivers of the regional epidemic. **In 1.5**, we combine epidemiological, diagnostic and viral genetic data to determine transmission chains within the HH: if, when and how long does it take for other family members to become infected (especially children) and cluster analysis using viral WGS. The transmission of SARS-CoV-2 from/to the community by HCW and their family members is being addressed here.

In **WP2** we strive to describe the spectrum of acute clinical presentation and the interrelation with paraclinical parameters in COVID-19 infected individuals that do not merit or get admitted to hospital. As we have enrolled HH members of COVID-19 positive individuals (see Methods), we are able to identify the whole spectrum of disease including asymptomatic individuals. Clinical phenotypes for hospitalised patients have been described in recent studies [23, 35]. However, mild COVID-19 disease presentation has been classified in broad categories [19]. **In 2.1** we attempt to define clinical phenotypes combining clinical and laboratory parameters collected in a systematic manner during the acute disease and correlate these with prognosis. Potential (risk) factors for mild versus more severe clinical evolution will include age, gender, BMI and specific blood parameters that have been related with severe disease progression such as C-reactive protein, hepatic transaminases, lactate dehydrogenase (LDH), leptin, interleukin (IL)-6, ferritin & other [36].

Clinical evidence on the subacute and long-term persistence of symptoms after SARS-CoV-2 infection or Long COVID, was recognised during the first few months of the COVID-19 pandemic [37, 38]. The persistence of symptoms, similar to myalgic encephalomyelitis/chronic fatigue syndrome, likely result from complex mechanisms involving an abnormal immune response to infection, central and autonomic nervous system and metabolic dysfunction among other mechanisms [39]. Viral infections are important triggers and may lead to what is often referred to as post-viral fatigue syndrome (PVFS) or post-covid fatigue syndrome, in cases that follow SARS CoV-2 infection. In COVID HOME, we follow consenting individuals at 3, 6, 12 and 18 months post diagnosis to evaluate fatigue and related symptoms. The data collected in **2.1** is used in **2.2** to identify clinical and laboratory parameters (biochemical and immunological), and clinical phenotypes, associated with the development of long-term sequelae. Immunopathology appears to play a very important role in SARS-CoV-2 pathogenesis. Critical disease has been found to be associated with a ‘cytokine storm’, the overproduction of inflammatory cytokines, ultimately resulting in acute respiratory distress syndrome (ARDS) [40]. In hospitalised patients, elevated levels of various cytokines, including IL-1β, IL-6, IL-10, IL-12, and tumor necrosis factor (TNF)-α, and chemokines, in particular CXCL10 and CCL2, have been found and the levels partly correlated with disease severity [41]. Cytokines may also play an important role in less severe disease courses and in Long COVID condition [42]. In **2.3** the dynamics of these cytokines will be determined during the acute disease and the long-term clinical evolution. Finally, in **2.4** we test the relationship of viral load dynamics and anti-SARS-CoV-2 antibody kinetics (collected in WP1 sections 1.1-1.3) with the clinical evolution. Viral loads of asymptomatic individuals have been reported to be similar to symptomatic patients [43]. However, in the case of anti-SARS-CoV-2 antibodies, reports suggest a weak immune response in mild clinical cases, as mentioned above. We aim to generate a larger set of data in relation to asymptomatic, mild and more severe disease presentation to answer these questions.

## Methods

### Study population and study area

The study is carried out amongst residents of the Northern Provinces of the Netherlands (Groningen, Friesland, Overijssel and Drenthe) comprising around 1.7 million inhabitants. Laboratory confirmed SARS-CoV-2 infected individuals of all ages (index cases) that do not merit hospitalisation, and their HH members, were invited to join the study and enrolled after signing a written informed consent. Consenting parents/guardians of infected children (≤16 years) signed the informed consent form. The complete HH, i.e. all persons (contacts) living in the same home, was to join the study at enrolment. If this requirement was not fulfilled, the HH was excluded from the study.

### Study design

An ongoing prospective longitudinal observational study of SARS-CoV-2 infected individuals that did not require hospitalisation and their HH members was initiated on March 2020, at the start of the pandemic in the Netherlands.

Enrolled individuals comprised the identified laboratory COVID-19 index positive patients plus all their HH members. The average number of HH inhabitants in the Netherlands and the northern region is 2.14 [44], thus for each enrolled patient we expected to enrol an average of 1-2 HH members in addition. Patients were followed daily at home approximately during 3 weeks post-infection to characterize their acute clinical phenotype, and are currently being followed at 3, 6, 12 and 18 months to determine their long-term clinical and serological evolution. During the follow-up, clinical data and different types of samples were/are collected, as described below.

### Inclusion criteria of index participant

The index participant was the first laboratory-confirmed SARS-CoV-2 infected individual of any age identified in a HH and invited to join the study together with the complete HH. The requirements for inclusion were:

- Onset of symptoms <5 days before inclusion
- Positive diagnostic RT-PCR for SARS-CoV-2 performed within 48h of inclusion
- Index person should supply all required samples in the protocol for the duration of the study
- All HH members should join the study
- Signing of informed consent form

### Data and sample collection

#### a. Identification of index cases

Potential index cases were identified at the virology facility of the Department of Medical Microbiology and Infection Prevention (MMBI, UMCG) and at the laboratories reporting to the GGD of the Northern Provinces. Suspected SARS-CoV-2 infected individuals were diagnosed by real-time RT-PCR performed on material from nasopharyngeal/throat (NPT) swabs. Laboratory SARS-CoV-2 positive individuals were contacted by phone by an infection prevention expert from either the UMCG MMBI department or the GGD who explained the study objectives and procedures and invited the positive individual and the HH members into the study if the inclusion criteria were met. Eligible individuals were also directed to the project’s website (https://www.umcg.nl/-/covid-home) for further information. Patients were also given time to ask questions and/or to think before deciding to join.

#### b. First home visit

Once a patient agreed to participate in the study, a first home visit (from now on called Day 0) was planned within the next 24 hours to enrol participants into the study after signing a written informed consent corresponding to the participant’s age (in duplicate, dated and signed by both participant and project researcher). Participants were asked to supply their individual e-mail address in the informed consent and to authorise it’s use to receive electronic questionnaires and information from the COVID HOME researchers via the project’s email. Participants were also encouraged to use the project’s email to ask questions or to inform of any event. A schedule was drawn so that study participants were visited weekly for at least 21 days by project staff to collect data and samples according to the different protocols. Those not tested or who were negative for SARS-CoV-2 RT-PCR, were enrolled into the protocol for HH members (section d).

#### c. Follow-up of SARS-CoV-2 infected individuals (acute phase of the disease/baseline)

A systematic weekly follow-up at the patient’s home was set up to acquire data and samples to determine kinetics, length, viability and routes of viral shedding (objectives 1.1 & 1.2), serological dynamics (objective 1.3), cytokine kinetics (objective 2.3) and evolution of haematological & biochemical blood parameters and their relationship with the patient’s clinical progression (objectives 2.1, 2.2, 2.4).

Enrolled patients were visited at home soon after initial diagnosis (Day 0), and then weekly on days 7, 14 and 21 after confirmed infection (Fig 1) to obtain clinical data and a blood sample for laboratory parameters, cytokine and serological determination. Research staff also obtained an NPT swab, urine and stool samples; and sperm or vaginal secretion (if consenting) from adult individuals (>16 years) to test for SARS-CoV-2 by RT-PCR (viral shedding) and for viral culturing (Fig 2). Children ≤16 years were only required to supply an NPT swab, urine and stool samples. Different paper and electronic case report forms (CRF/eCRF) were applied as described below (section d.) and in Fig 3. PCR results were communicated to the participants the following working day. If still positive on Day 21 for any of the specimens, the participant was invited to continue weekly sampling for RT-PCR testing for two further weeks.

**Fig 1.**
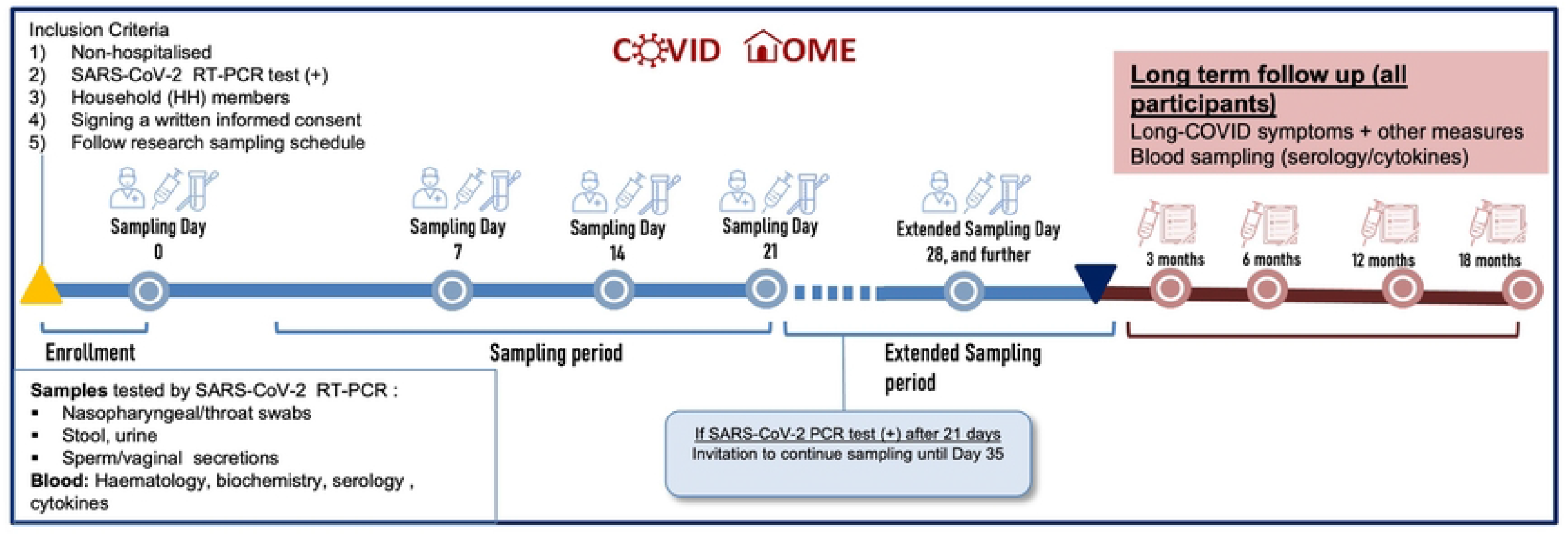
Acute and long-term follow-up timeline of participants enrolled in the COVID HOME study.

**Fig 2.**
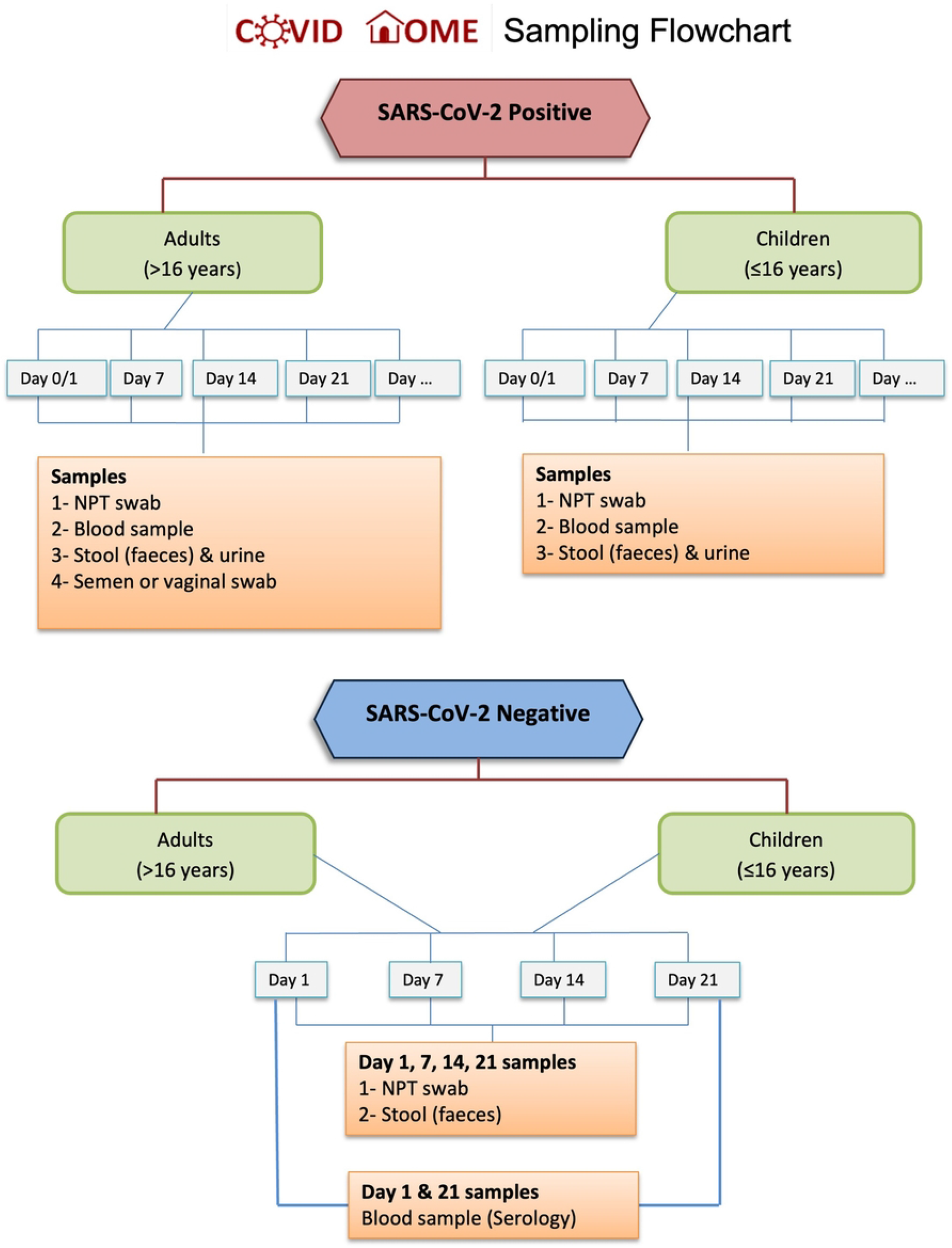
Sampling flowchart for positive and negative SARS-CoV-2 participants. NPT= nasopharyngeal/throat

**Fig 3.**
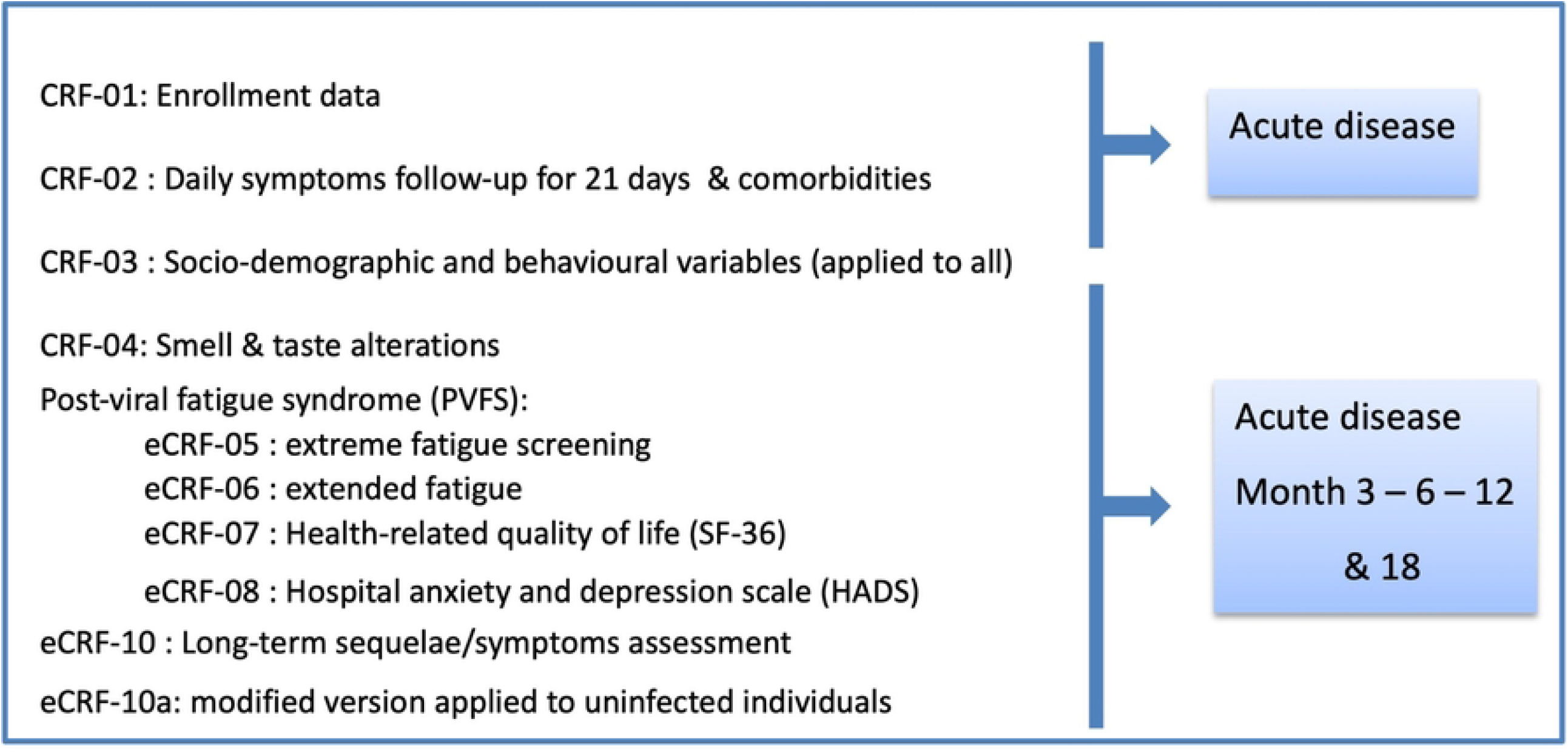
Case report forms (CRF) applied to study participants. CRF-01 and CRF-03 are applied to all participants while CRF-10a is answered by non-infected individuals. All other CRFs are applied to SARS-CoV-2 infected individuals.

Research staff visiting infected individuals at home followed safety measure protocols designed according to UMCG and national guidelines. Standard operating procedures (SOP) including flow-charts were designed for: i) safety procedures during home visits and at work, ii) handing/sending and retrieving CRFs from participants, iii) preparation of sampling materials and transport requirements, iv) sampling strategy (Fig 2), and v) aliquoting and storage conditions of samples; as well as protocols for sample processing.

#### d. Case report forms

The following CRF/eCRFs were/are applied to infected individuals (Fig 3):

▪ A self-completing clinical questionnaire (CRF-02) was given to patients at enrolment to record socio-demographic data, comorbidities, treatment and daily clinical history (symptoms and signs) during at least 21 days, and further if symptoms were still present. Participants still negative by RT-PCR but presenting symptoms were encouraged to fill this form as well.
▪ An individual HH questionnaire (CRF-03) was applied at enrolment to record socioeconomic characteristics, preventive measures, behaviour and intradomiciliary isolation measures taken.
▪ A phone interview (CRF-04) was carried out at 21 days, and at 3 and 6 months to assess smell and taste alterations in detail in consenting individuals.
▪ To evaluate the development of PVFS as part of the assessment of Long COVID, participants older than 16 years were asked to answer on Day 14 and then at 3, 6, 12 and 18 months post-infection to:
  ▪ The fatigue screening questionnaire (eCRF-05) to establish presence or absence of fatigue/tiredness, length of time of fatigue/tiredness, and any loss of activity
  ▪ Additional fatigue questionnaire (eCRF-06) to identify presence and severity of symptoms associated with development of PVFS
  ▪ Health-related Quality of Life (RAND-36) questionnaire (eCRF-07) and to
  ▪ the Hospital Depression and Anxiety Scale (HADS) questionnaire (eCRF-08)
▪ At 3, 6, 12 and 18 months post-infection, both adults and children receive eCRF-10 to assess Long COVID related symptoms, new comorbidities and re-infection.

CRF-02 and CRF-03 were given to participants to fill in paper while the rest were eCRFs and were answered online using the REDCap™ (Research Electronic Data Capture) secure web-based software platform [45]. Automatic electronic reminders were set up. In case of failure to complete the eCRFs, participants were called to check if email addresses were correct or if any other issues had arisen.

#### e. Follow-up of HH members of enrolled individuals (baseline)

HH members who had not been tested or were negative for SARS-CoV-2 RT-PCR at enrolment were followed with weekly testing of NPT swabs and stool samples for 3 weeks to determine if and when each individual became infected. At the beginning of the study, it became apparent that some individuals, especially children, stayed RT-PCR negative for NPT swabs, but became positive for SARS-CoV-2 infection when stool samples were tested by RT-PCR. Therefore, both NPT swabs and stool samples were collected weekly from (still) negative individuals. This information is being used to understand transmission chains, timing of infection between close contacts (family/HH members) and family cluster dynamics (objective 1.5). Moreover, to ascertain if RT-PCR negative individuals had been previously infected or seroconvert during follow-up, blood samples were drawn at enrolment and on Day 21 for IgM and IgG serology testing.

All HH members were asked to fill CRF-03 at enrolment and CRF-02 if presenting symptoms regardless of RT-PCR results. Once an individual was confirmed by real-time RT-PCR of any of the samples (NPT swabs and/or stool), the protocol for COVID-19 infected individuals (section c.) was followed.

#### f. Long-term follow-up

At 3, 6, 12 and 18 months post-infection, SARS-CoV-2 positive participants are invited to answer PVFS and eCRF-10 questionnaires sent digitally to determine post-COVID long-term sequelae (section d.) and provide blood samples for serologic determination.

Likewise, individuals that remained RT-PCR negative for SARS-CoV-2 infection have a blood sample taken at 3, 6, 12 and 18 months after enrolment for serological testing and a slightly modified version of eCRF-10 (eCRF-10a) is applied to attempt to differentiate long-term symptoms associated to COVID-19 from other causes, such as lockdown measures.

#### g. Substudy on smell and taste alterations

Smell and taste alterations are common in the acute phase during COVID-19 infection [46]. Alterations in smell and taste can have a profound impact on daily life and quality of life (QoL) by decreasing food liking and appetite [47]. However, the long-term impact has not yet been studied in COVID-19 patients. This substudy assesses the prevalence, characteristics and the impact of smell and taste alterations on daily life and QoL in COVID-19 patients at 3 weeks, and at 3 and 6 months after diagnosis. Knowledge on these alterations could lead to more personalized guidance of COVID-19 patients by their physician or a dietician.

Consenting study participants were called three weeks after COVID-19 diagnosis to fill in a questionnaire (CRF-04) regarding smell and taste alterations by phone. The questionnaire was based on a previously used questionnaire which was designed to study taste and smell disturbances in patients with gastrointestinal stromal tumors treated with tyrosine-kinase inhibitors [48]. The questionnaire was adjusted to study COVID-19 patients. It contains four parts where participants report the characteristics and impact of smell and taste alterations. The first and second part was completed by all participants, the third and fourth part was completed only by patients who reported changes in smell and taste respectively.

### Laboratory data and sample processing

Protocols and SOPs for sample processing were designed and updated according to the evolving knowledge on SARS-CoV-2 diagnosis and clinical evolution.

#### SARS-CoV-2 real-time quantitative Reverse Transcription PCR (qRT-PCR)

Viral load kinetics and duration of viral shedding of SARS-CoV-2 RNA in bodily specimens of all origins (respiratory, faeces, vaginal swab, urine, semen) were determined using an in-house protocol (in-house qPCR) [49]. Nucleic acids were extracted from 190µl of sample material, in addition to 10µl of internal control (phocine distemper virus (PDV)), using the NucliSense EasyMag or eMAG with the Specific A protocol (bioMerieux, Lyon, France), according to the manufacturer’s instructions. The in-house qPCR targets the SARS-CoV-2 E gene following the description by Corman *et al* [50] with minor modifications. Analysis is performed using a middleware software referred to as FlowG (LabHelp Labautomation), which can be used for interpretation of the results and communicating them back to the laboratory information system. Samples with a cycle time value (Ct) lower than 34 are considered positive while a Ct of 34-39 is considered inconclusive and the sample is repeated, which is routine clinical practice in our hospital for any new cases. Finally, Ct values above 40 are considered negative. The protocol was adapted to target new SARS-CoV-2 viral variants.

#### SARS-CoV-2 virus isolation and culture

The presence of infectious virus in different SARS-CoV-2 positive samples (nasopharyngeal/ throat, stool, urine, sperm/vaginal specimens) was ascertained through virus isolation in the BSLII facility of the UMCG. Samples cleared were spun down and filtered through 0.2 µm filter and characterised by a confirmatory RT-PCR positive test for COVID-19 and negative for a panel of other respiratory viruses. The presence of infectious SARS-CoV-2 virus was monitored observing for cytopathic effect (CPE) according to protocol for 7 days, and at day 2 and 7 using an in-house flow cytometry-based assay detecting viral antigen. Samples were considered positive for the presence of infectious SARS-CoV-2 virus if the flow cytometry-based assay showed more than two standard deviations from the mock-infected cells. Samples scoring positive in CPE but negative in flow cytometry-based assay were considered negative.

#### Anti-SARS-CoV-2 IgM, and IgG antibodies

Serological measurements (objective 1.3) are performed on the Abbott Architect platform, using a chemiluminescent magnetic microparticle immunoassay performed at the virology facility of the UMCG. Samples obtained during the acute disease were tested with the Semi-quantitative Architect IgM (AdviseDx SARS-CoV-2 IgMSP assay), and Semi-quantitative Architect IgG (Abbott Alinity i SARS-CoV-2 anti-nucleocapsid protein IgG assay), while long-term samples at 1, 3, 6, 12 & 18 months are tested with the Quantitative Architect IgG (AdviseDx SARS-CoV-2 IgG II) [51]. These results will be aligned per patient with the RT-PCR results, to evaluate the mean time points of IgM and IgG seroconversion, the range of IgM and IgG titres, and if seroconversion is related to viral clearance. We will determine the relation between Ct-values, viability of the virus and seroconversion. Patients are followed up to 18 months post-infection to assess the potential waning of immunity, the effect of vaccination and potential re-infections. The results of the serological testing are sent to the participants by post.

#### SARS-CoV2 whole-genome sequencing

To fulfil objective 1.4, nucleic acid extraction, purification and concentration from positive samples was performed after which, cDNA was prepared using a sequence-independent single-primer amplification (SISPA) approach, based on the round A/B methodology. Amplified cDNA was purified using a 1:0.8 ratio of AMPure XP beads (Beckman Coulter, Brea, CA), and quantified using a Qubit high-sensitivity double-stranded DNA (dsDNA) kit (Thermo Fisher), both according to the manufacturers’ instructions. Multiplex sequencing libraries was prepared using 250 ng of cDNA from up to 5 samples as input to the SQK-LSK109 and barcoded individually using the EXP-NBD104 Native barcodes (Oxford Nanopore Technologies). Libraries were sequenced on FLO-MIN106 flow cells on the MinION Mk1b device (Oxford Nanopore Technologies), with sequencing proceeding for up to 24 h.

#### Phylogenetic analysis to determine viral diversity and potential chains of transmission (Objectives 1.4 & 1.5)

Consensus sequences will be aligned with relevant sequences from SARS-CoV-2 lineages using MAFFT v7.313 [52]. Maximum likelihood phylogenies will be estimated using RAxML v8.2.12 (Stamatakis 2014) under a GTR-CAT nucleotide substitution. Viral lineage diversity and transmission clusters will be evaluated from the estimated phylogenies.

#### Haematological and biochemical blood parameters

As part of objectives 2.1-2.4, blood samples were processed at the routine UMCG laboratory to determine full blood count, haematocrit, C-reactive protein, creatinine, glycaemia, albumin, hepatic transaminases (AST, ALT), bilirubin, creatine kinase (CK), CK-MB isoenzyme, LDH, Prothrombin time (PT-INR), troponin, ferritin, procalcitonin, D-dimer, leptin and IL-6.

#### Cytokine/chemokine profiling

A panel of cytokines and chemokines (including IL-1β, IL-6, IL-10, IL-12, and TNFα, and chemokines, in particular CXCL10 and CCL2) is being measured using Luminex-based multiplex technology. Serum samples and associated clinical and Luminex/ELISA data will be combined and shared between different UMCG-COVID-19 research teams working on complementary studies addressing different COVID-19-related research questions. These markers will be used to assess prediction of disease severity and of the development of long-term symptoms.

### Data analysis

Due to the exploratory and mostly descriptive nature of the analysis, we did not conduct an exact sample size calculation. However, we focused on the prevalence of long-term sequelae as initial observations indicated that close to half of the infected individuals continue to suffer from symptoms after 4 weeks from infection [53, 54]. Therefore, we estimated that if the prevalence of Long COVID in the outpatient infected population was 40%, then a sample size of 256 individuals would give 80% power to estimate this prevalence with a +/-3% margin of error. Given the intense follow-up, we expected to enrol a minimum of 275 individuals, accounting for a 10% loss to follow-up, for a final sample size of 250 participants. Data is analysed anonymously using STATA (Stata Statistical Software: Release 14. College Station, TX,), R (version 3.6.3) and/or SPSS (IBM SPSS Statistics 23 (IBM Corp., Armonk, NY)) softwares.

#### Acute disease

Clinical presentation during the acute disease will be firstly presented in a descriptive analysis where the systematic prospective data collection will be used to better characterise mild clinical cases. We will attempt to define clinical phenotypes as endpoints and factors related to each phenotype. Principal component analysis-based cluster analysis will be used to group relevant variables of interest such as age, gender, comorbidities, signs, symptoms, haematological and biochemical parameters into phenotypes. Multinomial logistic regression models will be fitted to estimate associations of phenotypes with exposure variables such as viral loads, genetic sequence of SARS-CoV-2, anti-SARS-CoV-2 antibody kinetics, laboratory parameters and cytokine/chemokine levels. Clinical phenotypes will be categorised from milder to more severe disease presentation and potential predicting factors (measured at presentation) will be identified using generalized linear mixed models.

Generalised mixed models will be also used to assess the associations between the duration and severity of signs/symptoms of COVID-19 disease and viral loads, anti-SARS-CoV-2 antibody kinetics and change in laboratory parameters and cytokines, after adjusting for age and gender. The likelihood ratio test will assess statistical significance of the estimates at the 5% level. Univariate and multivariate analyses of potential risk factors for disease severity (mild versus more severe) will be performed using logistic regression comparing crude and adjusted odds ratios. Variables with a P-value ≤ 0.2 after adjusting by age and gender will be fitted into multivariate logistic regression models and adjusted for further confounders. Effect modification will be analysed and resulting models compared by likelihood ratio test.

A generalised linear regression model (Poisson distribution and log-link function) will be fitted to analyse the relationship between the duration of viral/RNA shedding and the exposure variables (viral loads, genetic diversity, antibody titres, laboratory parameters). Significance will be determined at the 5% level.

Phylogenetic analysis will be performed to determine viral diversity and potential chains of transmission within HHs and into the community as explained under Rationale above.

#### Long COVID

The presence of long-term sequelae at 3,6,12 & 18 months will be defined as reporting any symptoms and/or not feeling fully recovered at each of these time-points, as well as assessing the health-related quality of life using the SF-36 score at the different time points. These three definitions will be used as endpoints. A descriptive analysis will be performed to compare symptom presentation between time points and symptoms characteristics, such as number of symptoms, type of symptoms, symptom presentation (recurrent, constant, or both), and severity. Univariate and multivariate analyses will be carried out to assess risk factors for Long COVID from relevant variables of the acute disease such as comorbidities, number and duration of symptoms, type of symptoms and socio-demographic variables of interest. Analysis strategy will be as described above.

## Ethics, data management plan and biobanking

This study has been approved by the Medical Ethical Review Committee of the UMCG (METc 2020/158) and follows international standards for the ethical conduct of research involving human subjects. All procedures employed in the clinical and related laboratory studies comply with national and European legislation in respect of research involving human subjects.

Informed consents including participant information sheets (available in Dutch and English) were obtained before the collection of participants’ data and samples. At enrolment, participants invited to join the study received a written and oral explanation of what the research entails. Adult informed consents were applied to individuals older than 16 years, while a parent/guardian informed consent was signed by the child’s parent/guardian and together with their child if aged between 12-16 years. Individuals may withdraw from the study or may not consent to parts of the study, without any consequences.

A detailed data management plan (DMP) has been prepared by the project data steward and approved by the principal investigator describing how data will be handled both during research and after the project is completed. The UMCG has a comprehensive ISO27001 certificate and is compliant covering all of its strategic activities: Healthcare, Research and Education. The UMCG Research Code was drawn based on this certification. The COVID HOME DMP complies with the UMCG quality policy, UMCG and University of Groningen (RUG) Research Code and national guidelines on ethical research.

All information obtained during the course of this study, including hospital records, personal data and research data will be kept strictly confidential according to the General Data Protection Regulation (GDPR) of the Netherlands. Data protection and privacy is coordinated by the data protection officer which supervises that research conducted within the UMCG is performed in accordance with the GDPR. Study data were/are collected and managed using REDCap electronic data capture tools hosted at UMCG. Electronic web-based eCRFs have been developed using REDCap (Research Electronic Data Capture) which is a secure, web-based software platform designed to support data capture for research studies [45]. Participants enter data online by means of an electronic data entry interface. Data collected in paper CRFs were entered by research staff into the REDCap eCRFs. Inventory of samples collected and related SOPs were/are recorded into eLABJournal (eLabNext, Bio-ITech BV), a secured electronic lab notebook for biobanking purposes available only to dedicated UMCG researchers. Clinical data and laboratory results were entered into electronic databases and merged for analysis. To protect privacy, all data and sample records are entered as pseudonymized records (using a study identification number), not allowing the personal identification of study participants. Identifiable information is linked to stored data or samples only by a protected Master list.

All data (including clinical, laboratory and viral genetic data) are stored in password protected databases and archived in institutional research drives secured under encryption by our UMCG Information and Communication Technology department that can only be accessed by authorized project personnel. Data storage is backed-up automatically every day on the servers of the UMCG ensuring data safety. Samples are stored and managed locally according to UMCG SOPs. Patient information sheets and informed consents are kept in a locked cabinet. The data will be stored at the UMCG for 15 years. Data that may be reported in scientific journals will not include any information that identifies the participants in this study. We follow a FAIR (Findable, Accessible, Interoperable and Re-useable) data policy. After an embargo period of approximately 12 months (subject to change) from the project culmination and publication of main results, the processed, pseudonymised data will be made available for re-use. Requests for re-use of data will be evaluated by the principal investigator, project manager and data steward who will check whether the research question falls within the scope of the informed consent. If data will be shared it will not include directly identifiable information and can only be (re-)used within the scope of the informed consent. Third party use of data is governed in part by intellectual property rules and agreements, which will become available after publication of the main results.

## Preliminary Results

A total of 276 participants belonging to 108 HH were enrolled in the study by the end of May 2021. Twenty (7.2%) were excluded since 16 had a false positive RT-PCR test result and 4 were uncompliant.

The final study population comprised 256 participants (103 HH), of which 30 (13 HH) were enrolled during the first COVID-19 wave (March-May 2020) and 226 (90 HH) during the second and third waves (October 2020 until May 2021). Of the final study population, 190 participants (74.2%) had a positive RT-PCR test result for SARS-CoV-2 by the end of the 21-day acute disease follow-up period. The proportion of SARS-CoV-2 infected individuals was fairly similar for both enrolment periods, 73.3% (n=22) for the first wave versus 74.3% (n=168) for the second and third waves. Most individuals (183/190, 96.3%) developed mild to moderate disease while 7 patients were hospitalised between 7-9 days post-infection and one of these died.

At the time of writing this manuscript, all participants had reached the 3 and 6 month time-points of the long-term follow-up with data and serological sampling being collected. However, due to budgetary constraints, the first wave participants were not included in the 3 month follow-up. Approximately 78% of participants have been sampled/data collected at 12 month and 23% at 18 month post-enrolment. Preliminary analysis has shown that 43% (52/121) positive individuals reported having complaints at 3 months post-infection, while 42.7% (61/143) had complaints at 6 months.

## Discussion

This study aims to close a knowledge gap on the impact and consequences of COVID-19 on non-hospitalised patients, in contrast to most studies that naturally, have concentrated on hospitalised and critically ill patients. Enrolment of participants was initiated soon after the first cases were detected in the Netherlands [5], however, the north of the country experienced a low incidence during the first wave of the pandemic. Therefore, most of the enrolment took place during the second and third waves (October 2020 – June 2021) of COVID-19 in the Netherlands [55].

Our COVID HOME study addresses key questions of relevance to guide patient care and inform public health guidelines for infection prevention and control in the community and HH situation in a single study. The enrolment of patients right after diagnosis will allow the early identification of parameters potentially indicative for progression to severe disease. This early identification cannot be achieved in hospitalised individuals who are usually admitted a week or more after disease onset. The parameters that will be collected in this study, such as viral load, antibody response, cytokine changes and clinical and laboratory parameters of patient evolution will consent the identification of independent factors associated with disease evolution, insight in other transmission routes, and immune response dynamics in non-hospitalised patients. Moreover, as HH members of COVID-19 positive individuals were also enrolled, we have been able to determine if other family members become infected (especially children), how long does it take to get infected and which are the transmission dynamics within the HH. It will be important to determine transmission chains between the HH and workplace of adults, especially for healthcare settings to inform biosafety guidelines. A better understanding of the clinical evolution of non-hospitalised patients and the factors influencing disease progress (viral, immunological and other) can be rapidly turned into “home standard of care” guidelines for patients in home isolation and their HH members, as well as shed light on potential new therapies and/or strategies preventing progression to severe COVID-19 disease. Determining the length of infectiousness of COVID-19 patients will either confirm current guidelines or prompt their change in relation to quarantine and isolation measures, contributing to limit the spread of COVID-19 in the Netherlands and elsewhere. Hence the importance of the collaboration with public health institutions (National Institute for Public Health and the Environment [RIVM]/GGD) and policy makers.

In the Netherlands, the first indications of long-term consequences of SARS-CoV-2 infections (what is now known as Long COVID or Post-COVID syndrome) appeared though a survey carried out by the Dutch Lung Foundation in June 2020 [37]. We have found a high prevalence of SARS-CoV-2 infected individuals reporting still having complaints at 3 and 6 months post-infection, similar to those found in hospitalised patients [56]. However, there is uncertainty about the prevalence of Long COVID, the risk factors for this condition in adults and children and the role of new variants. A recent review on post-acute COVID-19 patients [38] highlight the importance of understanding the long-term evolution of these patients and the identification of factors that could predict Long COVID manifestations. The COVID HOME study is following participants up to 18 months after enrolment in order to contribute knowledge on the impact of this incapacitating condition at individual and societal level.

## Data Availability

All relevant data are within the manuscript and its Supporting Information files.

## Acknowledgements

We would like to firstly thank all the participants of the COVID HOME study for their ongoing contribution to the project and to the understanding of COVID-19. Our thanks also go to the Clinical Virology (Molecular and Serology) unit and to the Infection Prevention section, in special to Jet van der Weerd (Head of Infection Prevention), all part of the Department of Medical Microbiology & Infection Prevention, UMCG; as well as Lars van Heerden (manager) and the staff from the Laboratory of General Haematology and Chemistry, Laboratory Medicine, UMCG; and to Myke Mol and Paul Koenes from the Service Desk Clinical Research Office, UMCG, the Netherlands. Lastly, we would like to thank the collaboration of the Municipal Public Health Services (GGD) of Groningen and Drenthe with the recruitment of study participants in this region.

## Notes

**Funding** The project received funding from the Netherlands Organisation for Health Research and Development (ZonMw) [1], grant 10430012010023, and as partner of the ORCHESTRA project [2] which has received funding from the European Union’s Horizon 2020 research and innovation programme under grant agreement No 101016167. The views expressed in this publication are the sole responsibility of the author and the Commission is not responsible for any use that may be made of the information it contains. The funders had and will not have a role in study design, data collection and analysis, decision to publish, or preparation of the manuscript.

### Competing Interest Statement

The authors have declared no competing interest.

### Funding Statement

AT, grant 10430012010023, Netherlands Organisation for Health Research and Development (ZonMw), https://www.zonmw.nl/nl/over-zonmw/ AWF, 101016167, European Union’s Horizon 2020 research and innovation programme, https://web-staging.orchestra-cohort.eu/ The funders had and will not have a role in study design, data collection and analysis, decision to publish, or preparation of the manuscript.

### Author Declarations

Date 7 July 2020 METc number METc 2020/158 NEW Title Prospective cohort study of non-hospitalised COVID-19 patients: determining length of isolation and patient clinical development at home (COVID-HOME study). Old title: UMCG COVID-19 patient follow-up study: Understanding SARS-Cov2 infection and patient clinical development. UMCG RR number 202000198 The Medical Ethics Review Board of the University Medical Center Groningen (METc UMCG) has discussed the amendment regarding the above mentioned protocol. The committee considered whether or not the proposed changes lead to a different outcome. Based on the submitted documents the METc UMCG concludes that the amended protocol does not change the fact that the trial does not fall within the scope of the WMO. It is still not a clinical research with human subjects as meant in the Medical Research Involving Human Subjects Act (WMO). Kind regards, the Medical Ethics Review Board

